# Can machine learning or deep learning discover novel signatures of illness in continuous cardiorespiratory monitoring data?

**DOI:** 10.1101/2024.02.03.24302230

**Authors:** Brynne A. Sullivan, Ian G. Mesner, Justin Niestroy, Douglas E. Lake, Karen D. Fairchild, J. Randall Moorman

## Abstract

**Background:** Cardiorespiratory deterioration due to sepsis is a leading cause of morbidity and mortality for extremely premature infants with very low birth weight (VLBW, birthweight <1500g). Abnormal heart rate (HR) patterns precede the clinical diagnosis of late-onset sepsis in this population. Decades ago, clinicians recognized a pattern of reduced HR variability and increased HR decelerations in electrocardiogram tracings of septic preterm infants. A predictive logistic regression model was developed from this finding using mathematical algorithms that detect this signature of illness. Display of this model as the fold increase in risk of imminent sepsis reduced mortality in a large randomized trial. Here, we sought to determine if machine learning or deep learning approaches would identify this uncommon but distinctive signature of sepsis in VLBW infants.

**Methods:** We studied VLBW infants admitted from 2012 to 2021 to a regional Level IV NICU. We collected one-hour HR time series data from bedside monitoring sampled at 0.5 Hz (n=300 HR values per series) throughout the NICU admission. First, we applied the principles of highly comparative time series analysis (HCTSA) to generate many mathematical time series features and combined them in a machine learning model. Next, we used deep learning in the form of a convolutional neural network on the raw data to learn the HR features. The output was a set of HR records determined by HCTSA or deep learning to be at high risk for imminent sepsis.

**Results:** We analyzed data from 566 infants with 61 episodes of sepsis. HCTSA and deep learning models predicted sepsis with high out-of-sample validation metrics. The riskiest records determined by both approaches demonstrated the previously identified HR signatures-reduced variability and increased decelerations.

**Conclusions:** We tested the ability of unguided machine learning approaches to detect the novel HR signature of sepsis in VLBW infants previously identified by human experts. Our main finding is that the computerized approach returned the same result - it identified heart rate characteristics of reduced variability and transient decelerations. We conclude that unguided machine learning can be as effective as human experts in identifying even a very rare phenotype in clinical data.

## Introduction

Modern artificial intelligence approaches to physiological time series analysis have taken several worthy and productive forms. First, there is a round-up-the-usual-suspects approach with toolboxes of well-established analytics in the time-, frequency-, and non-linear dynamical domains (1). Second, there are analytics that are tailor-made for specific, clinician-identified abnormalities that make pathophysiological sense (like the paradoxical pulse of cardiac tamponade) or have stood the test of time (like ST-segment elevation in acute myocardial infarction).

In 2013, a novel approach was put forward by Fulcher and coworkers. They applied thousands of time series algorithms to many time series examples of all kinds (2). This highly comparative time-series analysis is not limited by the imagination nor by the knowledge or experience of the investigator and thereby allows the possibility of true discovery. It is, though, indiscriminate in nature. Algorithms that address specific problems in one domain (say, economics), are applied to specific clinical problems of an entirely different nature (say, sepsis). This allows the possibility of random associations with no robust, interpretable foundation for future applications.

Finally, there is the unsupervised approach of deep learning methods such as convolutional neural networks, where the computer is tasked with recognizing patterns more likely to occur when the outcome is present. We applied the principles of highly comparative time series analysis to the common and dire clinical problem of sepsis in premature infants. Previously, we identified a unique time-series signature of illness - abnormal heart rate characteristics of reduced variability and transient decelerations (3). These findings are pathognomonic; they are not regularly present in any clinical setting other than in distressed fetuses and in premature infants early in the course of sepsis. The signature was identified by a gold standard approach, namely domain experts who were highly familiar with the nuances of the clinical scenarios and with the phenotypes of normal cardiovascular regulation looked at continuous heart rate data from many infants in sickness and in health. The finding of low variability of HR punctuated by repetitive transient decelerations being associated with impending clinical deterioration from sepsis has clinical utility. Previously, we developed and optimized mathematical methods that were targeted to detect these abnormal heart rate characteristics (5,6). Display of a real-time heart rate characteristics score showing the risk of imminent sepsis (HeRO score) led to reduced deaths of VLBW infants in a very large multicenter randomized trial (4).

Recently, HR and oxygen saturation (SpO2) features have been associated with other adverse neonatal outcomes. An algorithm that detects a lack of successive HR increases predicts all-cause mortality (7), HR skewness predicts autism spectrum disorder (8), HR-SPO2 cross-correlation predicts sepsis and necrotizing enterocolitis (NEC) (9), and complex lagged-difference HR metrics predict cerebral palsy (10).

In this study, we sought to test the hypothesis that unguided approaches to discovery, HCTSA with machine learning and deep learning with neural networks, identify the same unique HR signature of sepsis in premature infants as was recognized by human experts. Proving this hypothesis will inform future work on feature discovery, where either human expert inspection of physiological data or unguided AI approaches can identify the same signatures of illness. Disproving our hypothesis by discovering novel features associated with neonatal sepsis will provide evidence for the utility of the unguided approaches.

## Methods

### Patients

We studied VLBW infants admitted to the University of Virginia Children’s Hospital NICU from 2012 to 2022. The study was approved by the Institutional Review Board at the University of Virginia with a waiver of informed consent. We excluded infants with major chromosomal or congenital anomalies and those with no vital sign data collected. We collected demographic and clinical variables from the electronic health record and unit databases.

### Sepsis definition

The primary outcome was late-onset sepsis (LOS), defined as a positive blood culture obtained beyond 3 days of age and treated for at least five days with antibiotics. We excluded blood cultures that were negative, those that were positive within seven days of a prior positive blood culture, and those treated as contaminants (defined as treated with <5 days of antibiotics). The time that a positive blood culture was ordered was used as the time of sepsis diagnosis.

### HR data collection and preprocessing

Continuous electrocardiogram-derived HR data were collected from standard NICU bedside monitors (GE, Philips) using the BedMaster system (Hillrom’s Medical Device Integration Solution, Chicago, IL), sampled from the waveform at 0.5 Hz. HR values representing incontrovertible artifact (zeros) were removed, and times with missing data were omitted, not imputed. We sub-sampled the raw data by taking four random one-hour HR records per day. Each on-hour record was labeled as “1” if sepsis diagnosis occurred in the subsequent 24 hours or “0” if it did not. Records falling within 72 hours of birth and seven days of sepsis diagnosis were excluded.

### HCTSA and machine learning methods

In previous studies, we developed methods to derive a subset of representative algorithms from the 7700+ HCTSA algorithms cataloged by Fulcher, et al. in 2017 (11) and applied those methods here. First, we implemented 4246 instances of 111 algorithms from 9 families: measures of distribution (n=212), correlation (290), entropy (216), time-series modeling (58), stationarity (217), symbolic transforms (689), trend analytics (225), biomedical (40), and others (298). After eliminating those with unusable results (such as missing values), we proceeded with the results of 2133 mathematical operations. We used an unsupervised clustering method based on mutual information measures and identified 14 clusters that accounted for more than 80% of the variance. We selected a single algorithm at or near the medoid to represent each cluster. We used the results of these 14 algorithmic operations for each one-hour HR record as predictors of sepsis in the next 24 hours using multivariable logistic regression modeling adjusted for repeated measures. We determined the records with the highest predicted probability of sepsis and inspected them by eye.

### Deep learning methods

We used the InceptionTime PyTorch (12) model distributed within the tsai package (13) (Oguiza 2023), with constructs such as the attribution maps and focal loss implementations from the fastai package (https://docs.fast.ai/). Architecture defaults were left unchanged, but learning rates and batch sizes were tuned ad-hoc for hardware and model performance. The fastai learner.fit_one_cycle implementation of the 1cycle policy of Smith and Topin (2018) was used with the Adam optimizer and the focal loss function for training.

We applied Gradient-Weighted Class Activation Mapping (Grad-CAM) (14) methods to the deep learning model to identify the features that contributed to the model’s predictions. Grad-CAM extends basic class activation maps by applying the activation functions from the forward pass as an ask to the gradients from the backward pass. In doing so, the method is able to highlight the inputs related to each class’s prediction for a single one-hour record.

A minimal InceptionTime architecture was used, modified from example usages only to 1) establish reasonable learning rates and batch sizes, 2) calculate the receiver operator characteristic area under the curve (AUC) as a validation metric, 3) choose a validation dataset for patients not seen during training, and 4) to use a focal loss function to address the large class imbalance. Then, a series of experiments were done in an attempt to increase predictive performance as measured by AUC. As an ad-hoc attempt to address issues of autocorrelation and temporal patterns with offsets, the same analysis was performed using the single-index difference of the vital sign time series. Longer 20m and 1hr samples were extracted from the source data to determine if longer samples would increase performance. Finally, the same folds for cross-validation were implemented to assess inter-fold performance variability and also to have a direct comparison to other regression-based models.

To assess the attribution and interpretability of the model, Grad-CAM attribution heatmaps were created and displayed as 1D heatmaps of vertical bars below the sample time series. The last convolutional layer was used to provide temporal patterns of attribution, and attribution scores (derived from gradients) were min-max normalized for a consistent scale. The one-hour records with the highest risk scores were plotted and visually inspected. Model scores (softmax of final layer for each class) were converted to percentiles, and equidistant octiles (0:1:0.125) were plotted to show samples across model scores.

## Results

### Patients

We analyzed data from 566 infants with 61 episodes of sepsis. Table 1 summarizes cohort characteristics overall and grouped by infants with and without one or more episodes of late-onset sepsis. Infants with sepsis were of lower gestational age and birthweight than those without. We analyzed 64,477 one-hour HR records, of which 185 occurred within 24 hours of sepsis diagnosis.

|  | All | Sepsis | No sepsis |
| --- | --- | --- | --- |
| Gestational age (weeks), mean, SD | 28.1 (3.1) | 26.8 (2.8 ) | 28.2 (3.1) |
| Birth weight (grams), mean, SD | 990 (294) | 874 (261) | 1003 (295) |
| Male sex, n (%) | 307 (52) | 33 (54) | 274 (52) |

### Signatures of sepsis identified by machine learning and deep learning

We inspected the records with the highest predicted probability of sepsis in both the HCTSA machine learning and deep learning models. The visual patterns of these records were the same as those identified by human experts-low HR variability with more transient HR decelerations (Figures 1 & 2).

**Figure 1.**
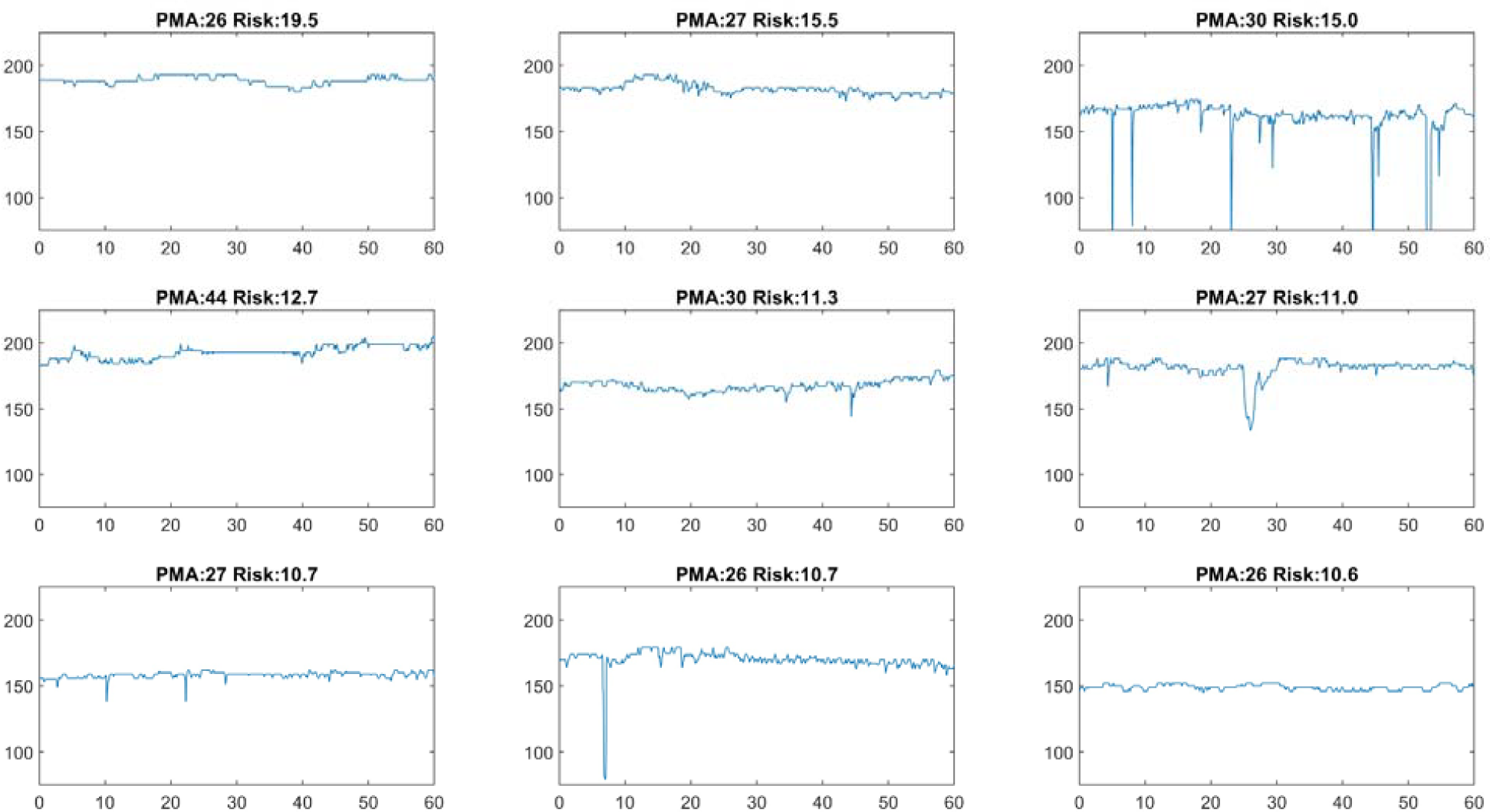
Nine heart rate records in the highest percentile of sepsis risk scores as predicted using HCTSA and machine learning. Visually, these time series have low variability with or without frequent, transient decelerations. PMA = post-menstrual age at the time of the record shown. Risk = the relative risk of sepsis in the next 24 hours.

In addition to expected measures of low variability, such as the coefficient of variation (15), the HCTSA machine learning model identified several novel metrics that are well-suited to detect neonatal HR time series with reduced variability and transient decelerations (Figure 1). These include:

1. Normalized nonlinear autocorrelation, in particular, the autocovariance at lag 1 (16).
2. Measures of the asymmetry above and below the mean; for example, record the minimum and the maximum value of non-overlapping windows and perform comparisons such as their differences and ratios. Here, the finding was a large value of the ratio of the medians of the local maxima and minima, corresponding to the presence of decelerations. In the original work, we developed sample asymmetry with the same goal in mind (5).
3. Symbolic logic, whereby ranges of values are assigned letters, and motifs are sought. Here, the finding was that only a small number of the possible motifs occurred frequently in the abnormal records, corresponding to the presence of reduced variability.

Deep learning using neural networks proved to be an unsupervised method for finding records of very low variability or frequent decelerations (Figure 2).

**Figure 2.**
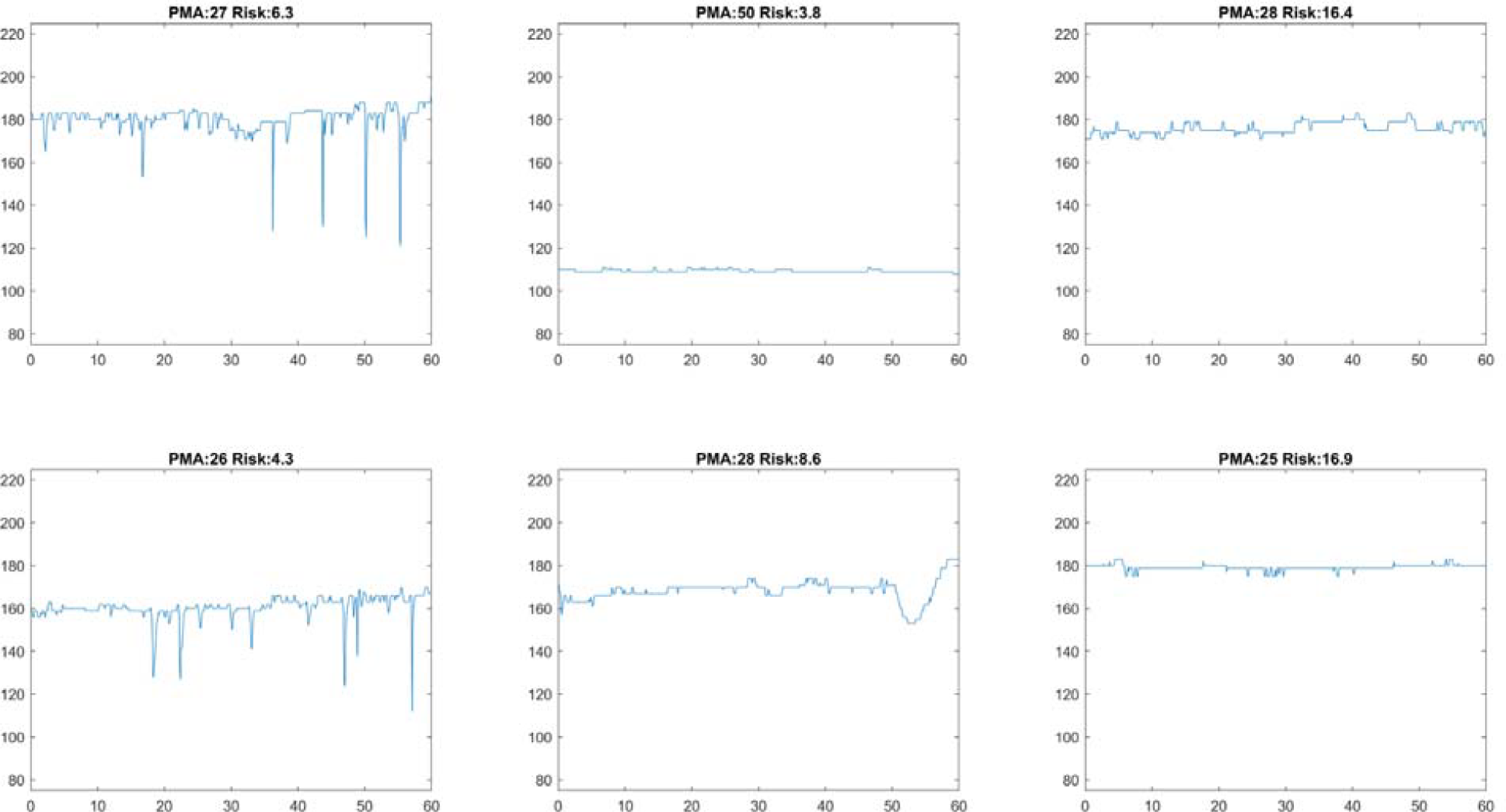
Six on-hour heart rate records in the highest percentile of sepsis risk scores as predicted using deep learning and activation mapping. Visually, these time series have low variability with or without frequent, transient decelerations. PMA = post-menstrual age at the time of the record shown. Risk = the relative risk of sepsis in the next 24 hours.

Our machine learning and deep learning models had a similar predictive performance as measured by out-of-sample AUCs (0.815 for the deep learning model, AUC 0.813 for the HCTSA model).

## Discussion

We used two data-driven approaches, machine learning with HCTSA and deep learning, to determine whether these methods would identify the same signatures of sepsis in premature infants as noticed by the human eye, or new ones. We found that both methods identified the highest risk records as those with low heart rate variability and transient heart rate decelerations-the same signatures identified by expert clinicians decades ago.

The signature of reduced variability and transient deceleration during neonatal sepsis was discovered by reviewing by hand many hundreds of HR time series near sepsis and during healthy periods. Once recognized, our group of experts in clinical neonatology and cardiology, statistics, and mathematical modeling set out to develop algorithms to detect this visual pattern (3,5,17). We tested the validity of these algorithms in the most rigorous way possible: the development of a predictive model on a large-representative cohort with external validation (18), followed by a multicenter, randomized clinical trial (4). This work demonstrated predictive validity across NICUs and clinical impact in the form of reduced mortality.

While this work was successful, it is inefficient and does not take advantage of modern AI methods for feature discovery. Thus, we sought to determine whether unguided machine learning approaches could do as well or better than visual inspection by human experts. We have applied HCTSA to physiologic time series data to discover signatures of conditions other than sepsis where the task of visual inspection had not been undertaken and would not be feasible. In this work, we found physiologic signatures of NICU mortality, cerebral palsy, and autism. These results have generated new hypotheses on the pathophysiology of the disease, but we could not prove that the same signature would have been identified had the researchers started with visual inspection of many physiologic time series. The historical research on abnormal heart rate characteristics in neonatal sepsis provided the ideal experiment to evaluate the utility of unguided machine learning for identifying physiological signatures of illness.

We attribute the favorable clinical impact of heart rate characteristics monitoring to its basis in physiology rather than the common practice of making predictive models based on electronic health records. Data on laboratory tests, medications, and interventions carry information that the clinical team already knows since they put in the orders. In contrast, signatures of illness in physiological monitoring data carry information from the patient that precedes clinical deterioration and therefore constitute reliable inputs to multivariable models of the future.

## Conclusions

Unguided machine learning carried out on heart rate time series identified the same physiologic signature of neonatal sepsis as recognized by human experts. This demonstrates the utility of such methods, which can be applied to other physiologic time series and patient populations toward discovering novel signatures of illness and building AI technology with clinical impact.

## Data Availability

All data produced in the present study are available upon reasonable request to the authors

## Bibliography

1. Vest AN, Da Poian G, Li Q, Liu C, Nemati S, Shah AJ, et al. An open source benchmarked toolbox for cardiovascular waveform and interval analysis. Physiol Meas. 2018 Oct 11;39(10):105004.

2. Fulcher BD, Little MA, Jones NS. Highly comparative time-series analysis: the empirical structure of time series and their methods. J R Soc Interface. 2013 Jun 6;10(83):20130048.

3. Griffin MP, Moorman JR. Toward the early diagnosis of neonatal sepsis and sepsis-like illness using novel heart rate analysis. Pediatrics. 2001 Jan;107(1):97–104.

4. Moorman JR, Carlo WA, Kattwinkel J, Schelonka RL, Porcelli PJ, Navarrete CT, et al. Mortality reduction by heart rate characteristic monitoring in very low birth weight neonates: a randomized trial. J Pediatr. 2011 Dec;159(6):900–6.e1.

5. Kovatchev BP, Farhy LS, Cao H, Griffin MP, Lake DE, Moorman JR. Sample asymmetry analysis of heart rate characteristics with application to neonatal sepsis and systemic inflammatory response syndrome. Pediatr Res. 2003 Dec;54(6):892–8.

6. Richman JS, Moorman JR. Physiological time-series analysis using approximate entropy and sample entropy. Am J Physiol Heart Circ Physiol. 2000 Jun;278(6):H2039–49.

7. Niestroy JC, Moorman JR, Levinson MA, Manir SA, Clark TW, Fairchild KD, et al. Discovery of signatures of fatal neonatal illness in vital signs using highly comparative time-series analysis. npj Digital Med. 2022 Jan 17;5(1):6.

8. Blackard KR, Krahn KN, Andris RT, Lake DE, Fairchild KD. Autism risk in neonatal intensive care unit patients associated with novel heart rate patterns. Pediatr Res. 2021 Dec;90(6):1186–92.

9. Fairchild KD, Lake DE, Kattwinkel J, Moorman JR, Bateman DA, Grieve PG, et al. Vital signs and their cross-correlation in sepsis and NEC: a study of 1,065 very-low-birth-weight infants in two NICUs. Pediatr Res. 2017 Feb;81(2):315–21.

10. Letzkus L, Picavia R, Lyons G, Brandberg J, Qiu J, Kausch S, et al. Heart rate patterns predicting cerebral palsy in preterm infants. Pediatr Res. 2023 Oct 27;

11. Fulcher BD, Jones NS. hctsa: A Computational Framework for Automated Time-Series Phenotyping Using Massive Feature Extraction. Cell Syst. 2017 Nov 22;5(5):527–531.e3.

12. Paszke A, Gross S, Massa F, Lerer A, Bradbury J, Chanan G, et al. PyTorch: An Imperative Style, High-Performance Deep Learning Library. arXiv. 2019 Dec 3;

13. GitHub - timeseriesAI/tsai: Time series Timeseries Deep Learning Machine Learning Pytorch fastai | State-of-the-art Deep Learning library for Time Series and Sequences in Pytorch / fastai [Internet]. [Cited 2024 Jan 30]. Available from: https://github.com/timeseriesAI/tsai

14. Selvaraju RR, Cogswell M, Das A, Vedantam R, Parikh D, Batra D. Grad-CAM: Visual explanations from deep networks via gradient-based localization. Proceedings of 2017 IEEE International Conference on Computer Vision (ICCV). IEEE; 2017. p. 618–26.

15. Griffin MP, Scollan DF, Moorman JR. The dynamic range of neonatal heart rate variability. J Cardiovasc Electrophysiol. 1994 Feb;5(2):112–24.

16. Schreiber T, Schmitz A. Discrimination power of measures for nonlinearity in a time series. Phys Rev E. 1997 May 1;55(5):5443–7.

17. Lake DE, Richman JS, Griffin MP, Moorman JR. Sample entropy analysis of neonatal heart rate variability. Am J Physiol Regul Integr Comp Physiol. 2002 Sep;283(3):R789–97.

18. Griffin MP, O’Shea TM, Bissonette EA, Harrell FE, Lake DE, Moorman JR. Abnormal heart rate characteristics preceding neonatal sepsis and sepsis-like illness. Pediatr Res. 2003 Jun;53(6):920–6.

